# Economic implications of polypharmacy in Nepal: Multicenter community-based study

**DOI:** 10.1101/2020.09.12.20193458

**Authors:** Binaya Sapkota, Suraj Shakya, Saroj Sah, Narendra Chaudhary, Bijendra Tamang, Saugat Singh Karki

**Affiliations:** Nobel College Faculty of Health Sciences, Sinamangal, Kathmandu, Pokhara University, Nepal.

**Keywords:** Adverse effects, community, economic implication, Nepal, polypharmacy

## Abstract

**Objective:** Improper management of polypharmacy in community and hospital pharmacies may lead to adverse effects and drug interactions in patients of all age groups, especially children and the elderly. This study aimed to determine the scenario of polypharmacy in multiple communities in Nepal and the costs associated with them.

**Design:** Cross-sectional study

**Setting:** Local communities of nine districts in Nepal

**Participants:** Total 400 patients of all age groups, who were consuming medicines and who fulfilled inclusion criteria from May 2017 to August 2018

**Primary and secondary outcome measures:** A semi-structured questionnaire, based on the prescription optimization method, was used for data collection. Multinomial logistic regression was performed to analyze the statistical significance of polypharmacy with the predictor variables (e.g., age, education level, occupation, diagnosis, total cost of medicines). The p-value < 0.05 was considered statistically significant at 95% confidence interval. Polypharmacy cases and their economic implications were reported.

**Results:** Eighty-one patients (20.3%) with an age group of 22-31 years with female patients (219, 54.8%) reported more polypharmacy events. There were 216 patients (54%) with prescriptions of five medicines i.e., moderate polypharmacy. Total number of medicines consumed by all 400 patients was 2269, with a mean±SD 5.67 ±1.08. Total expenditure by all 400 patients was USD 3409.54 during the study period, with a mean±SD8.66±6.04. Both moderate and severe polypharmacy cases were non-significantly related with age, gender and total cost of medications. They had significant relationship in almost all levels of education and occupation and showed mixed type of significance and non-significance with the diagnosis of the respondents.

**Conclusion:** Polypharmacy cases can be minimized, considering adverse drug reactions and drug interactions. Further studies are warranted in medication utilization and avoidable polypharmacy along with detailed pharmacoeconomic evaluation.

## Background

Polypharmacy is not an entity with unanimously accepted definition.^1-3^ There are many viewpoints that the concomitant use of multiple (more than four) medications by a patientisregarded as polypharmacy.^4-8^ Even worse is the excessive polypharmacy with ≥10 medications.^3,4^ Prevalence of polypharmacy was 7% in the USAand 1.2% in Denmark.^3,9^ Problems of polypharmacy may be encountered in all health care settings, including inpatients, an outpatient unit, discharge patients, long-term care facilities (LTCFs) such as nursing homes.^6^ Polypharmacy cases are yet the under-researched domains in the developing countries, including Nepal. Major risk factors for polypharmacycan begrouped into demographic (eg, advancing age, education), health status (eg, depression, hypertension, anemia, asthma, angina, osteoarthritis, gout, diabetes mellitus), and access tohealth care(eg, health care visits, health insurance).^1^ Therapeutic errors of treating ADRs of a medicine as a separate diagnosis further potentiate polypharmacy.^6^

Polypharmacy poses a huge economic impact and significantly detriorates patients’ quality of life (QoL) via the potential therapeutic failure. In 2012, the US Institute for Healthcare Informatics found that preventable polypharmacy contributes to 4% of the costs that is equivalent to USD 18 billion worldwide. Incorporating pharmacoeconomic evaluations by the pharmacists along with the prescribers can reduce the health care burden posed by polypharmacy.^10^ Other complications such as adverse drug reactions (ADRs), interactions, non-adherence, and geriatric syndromes of organ dysfunction are also attributed to polypharmacy.^4,6-8,11^ Jansenand Brouwers concluded that pharmacists and geriatricians can play a pivotal role to optimize polypharmacy in the elderly.^6^ Polypharmacy casesare further associated withthe avoidable but significant increment of the cost of medications. Rozenfeld et al. studied medicines utilization by 800 retirees of the Brazilian Institute of Social Security aged ≥60 years with the average age 72.27 (64.5–80) years,and observed that muscle relaxants, antihistamines, and long-acting benzodiazepines led to anticholinergic adverse effects (eg, sedation, weakness, falls, and fractures).^12^

Research conducted byBasnet et al. in Nepal showed that 225 geriatric in-patients (aged ≥65 years) were prescribed with average of 1844 medicines during hospital stay, with an average of 8.19±3.5 per person.^13^ Similarly, Lohani et al. studied the prescribing pattern among geriatric in-patients (≥65 years) at Tribhuvan University Teaching Hospital (TUTH), Nepal from July 2002 to June 2003 and found an average of 10.73 medicines prescribed,89% of patients prescribed with > 5 medicines.^14^

## Aim of the study

The present study was intended to explorepolypharmacy with demographics and costs incurred with polypharmacy in select Nepalese communities.

## Methods and Materials

### Study design and study variables

Multi-center community-based cross-sectional study was conducted by collecting various predictor variables such as age, gender, education level, occupation, diagnosis,total cost of medications associated with polypharmacy as the known determinants of polypharmacy. Outcome variable such as status of polypharmacy based on number of simultaneous medications used was taken into consideration.

## Study area and study site

The study was conducted in local pharmacies of nine districts namely Kathmandu, Bhaktapur, Lalitpur, Nuwakot, Makawanpur, Chitwan, Dhanusha, Sunsari, and Siraha.The population distribution among the districts aforementioned was 435,544, 68,557, 109,505, 59,194, 86,045, 132,345, 138,225, 162,279 and 117,929 and altogether 1,309,623.^15^ The districts and study sites (ie, local pharmacies) were randomly selected (with the help of computer-generated table of random numbers) using the principles of multistage sampling, and finally simple random sampling technique.

## Patient and Public Involvement

The study includedthe uneducated andthe educated patients of all age groups, who were consuming medicines for their health problems. All the respondents who fulfilled the inclusion criteria within the study duration of May 2017 to August 2018 wereselected for the study.Respondents were selected based on the pharmacy exit interview.Patients’ exit interview was performed in the community pharmacies along with their prescriptions to avoid the potential bias.Data were collected for nine months, starting from May 2017, and one pharmacy for a month.

## Ethics approval

Ethics approval was taken from the Nobel College Institutional Review Committee (IRC), Sinamangal, Kathmandu (NIRC 0103/2017).National ethics guidelines developed by Nepal Health Research Council (NHRC) was followed during the whole research period. Both verbal and written informed consents were received from the respondents. However, the interview session was not electronically recorded, and was only manually recorded in the semi-structured questionnaire. In the case of vulnerable age group respondents, such as children under 16 years old, written informed consentwas obtained from their family guardians, and they were interviewed on their behalf.

## Inclusion and exclusion criteria

Respondents prescribed with polypharmacy were included in the study.Community people not willing to participate in the study, as well as those not taking medicines for their health problems, were excluded.

## Sample size

Altogether 400 patients from nine districts were selected based on the established formula for cross-sectional studies:

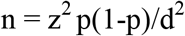

where, z = 1.96 at 95% confidence interval; p = 0.5 (since it was unknown); d = 0.05 Calculating the parameters, n = 385 (rounding to 400) was taken for the study.

## Data collection tools

Semi-structured data collection sheetbased on the prescription optimization method,developed for appropriate prescribing,^11^ was used for data collection.Researchers interviewed the respondents (originally in Nepali language and later transcribed in the English) along with a review of their prescriptions as well as over-the-counter (OTC) and prescription-only medications (POM) they were taking during the study period. The output of the interview was manually recorded in the data collection sheet. The sheetfilled were checked and initially processed (ie, edited, coded/decoded; later classified and tabulated during data analysis) at the field level on the same day of the data collection. Medicines prescribed were classified according to the ATC (Anatomic Therapeutic Chemical) classification proposed by the WHO.^16^(Annex 1 and Annex 2)Information about the cost of the medicines was taken from the maximum retail prices (MRPs) of the medications at the local pharmacies, where the study was performed. Categorization of the OTC or POM medications was based on the University of Maryland Health Partners (UMHP) OTC Drug List (updated on May 2018)^21^ and verified with the Nepalese Drug Act 1978.^17^

## Reliability and validity

Pretesting of the sheet was done in 10% of the total sample size estimated (ie, 40) in a similar setting as of the final study. The pretested data were utilized only for the modification of the sheet and were not included in the final data analysis. Internal validity (ie, confidence in concluding outcome due to the predictor variables) as well as external validity (ie, generalizability) were assured with the extensive literature review and timely pre-testing.

## Data analysis

Data were statistically analyzed with theR programming 4.0.1.^18^ Descriptive analysis wasperformed by mean, standard deviation, frequency distribution and percentage.Multinomial logistic regression analysis was performed to analyze the statistical significance status of polypharmacy with variouspredictor variables (eg, age, gender, education level, occupation, diagnosis, total cost of medicines). The p-value< 0.05 was considered statistically significant at 95%confidence interval. Status of polypharmacy was assessed as follows:mild polypharmacy: 1–4 medications, moderate polypharmacy: 5–9 medications, and severe polypharmacy: ≥10 medications.^19^ These all categorizations were based on the number of medications prescribed and/or consumed. Since the health system in Nepal is mainly focused on the out-of-pocket (OOP) payment, the cost of the medication was borne by the patients themselves.

## Results

Eighty-one patients (20.3%) aged 22-31 years, with more female patients (219,

54.8%)experienced polypharmacy. Two hundred twenty-three (55.8%) illiterate respondentsexperienced polypharmacy. More housewives (187, 46.8%) responded in the study. There were 204 patients (51%) from various locations of Siraha district. (Table 1) One hundred one patients (25.3%) reported problems of fever, cough, and muscle pain. (Table 2)There were 216 patients (54%) with prescriptions of five medicines. (Table 3)Total expenditure by all 400 patients was USD 3409.54, with mean±SD 8.66±6.04. (Table 4)

**Table 1:**
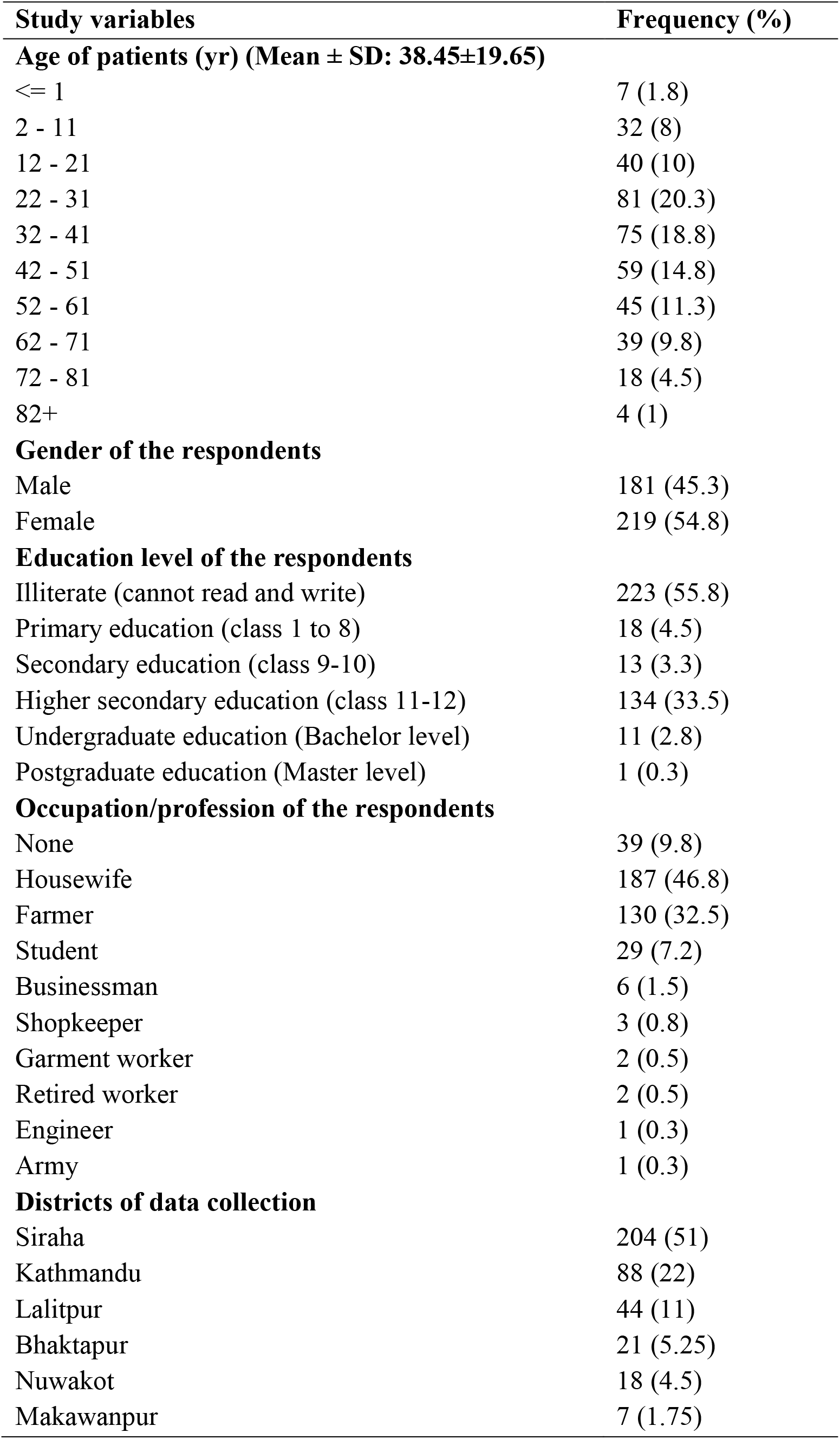
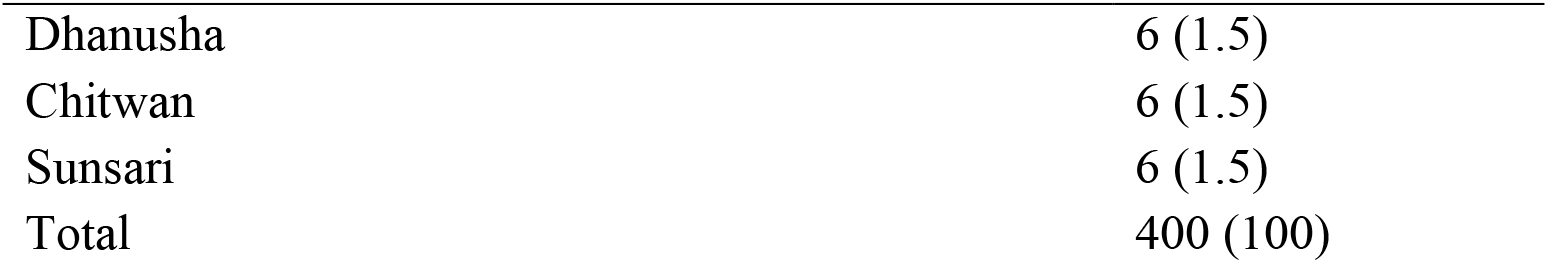
Demographic characteristics of study population (n = 400)

| **Study variables** | **Frequency (%)** |
| --- | --- |
| **Age of patients (yr) (Mean ± SD: 38.45±19.65)** |  |
| <= 1 | 7 (1.8) |
| 2 - 11 | 32 (8) |
| 12 - 21 | 40 (10) |
| 22 - 31 | 81 (20.3) |
| 32 - 41 | 75 (18.8) |
| 42 - 51 | 59 (14.8) |
| 52 - 61 | 45 (11.3) |
| 62 - 71 | 39 (9.8) |
| 72 - 81 | 18 (4.5) |
| 82+ | 4 (1) |
| **Gender of the respondents** |  |
| Male | 181 (45.3) |
| Female | 219 (54.8) |
| **Education level of the respondents** |  |
| Illiterate (cannot read and write) | 223 (55.8) |
| Primary education (class 1 to 8) | 18 (4.5) |
| Secondary education (class 9-10) | 13 (3.3) |
| Higher secondary education (class 11-12) | 134 (33.5) |
| Undergraduate education (Bachelor level) | 11 (2.8) |
| Postgraduate education (Master level) | 1 (0.3) |
| **Occupation/profession of the respondents** |  |
| None | 39 (9.8) |
| Housewife | 187 (46.8) |
| Farmer | 130 (32.5) |
| Student | 29 (7.2) |
| Businessman | 6 (1.5) |
| Shopkeeper | 3 (0.8) |
| Garment worker | 2 (0.5) |
| Retired worker | 2 (0.5) |
| Engineer | 1 (0.3) |
| Army | 1 (0.3) |
| **Districts of data collection** |  |
| Siraha | 204 (51) |
| Kathmandu | 88 (22) |
| Lalitpur | 44 (11) |
| Bhaktapur | 21 (5.25) |
| Nuwakot | 18 (4.5) |
| Makawanpur | 7 (1.75) |
| Dhanusha | 6 (1.5) |
| Chitwan | 6 (1.5) |
| Sunsari | 6 (1.5) |
| Total | 400 (100) |

**Table 2:** Diagnosis mentioned in the prescription of the respondents (n = 400)

| Diagnosis | Frequency (%) |
| --- | --- |
| Fever, Cough, Muscle Pain | 101 (25.3) |
| Hypertension, Diabetes mellitus, Gastritis, High Cholesterol level | 36 (9) |
| Kidney stone, Abdominal pain, Gastritis | 33 (8.3) |
| Muscle pain, Weakness | 23 (5.8) |
| Fever, Headache, Joint pain | 16 (4) |
| Constipation | 15 (3.8) |
| Cough, Fever, Headache, Tonsillitis | 15 (3.8) |
| Hyperthyroidism | 13 (3.3) |
| Nausea, Vomiting, Abdominal pain | 11 (2.8) |
| Type II diabetes | 11 (2.8) |
| Total<sup>#</sup> | 400 (100.5) |

<sup>#</sup>Only cases with frequency of two digits have been included in table.

**Table 3:** Number of medicines prescribed simultaneously and used by the patients (Mean ± SD: 5.67 ±1.083) (Total no. of medicines consumed by all 400 pts: 2269) (n = 400)

| No. of medicines | n (%) | Status of polypharmacy |
| --- | --- | --- |
| 4 | 16 (4) | Mild (1 to 4 medicines) (16, 4%) |
| 5 | 216 (54) | Moderate (4 to 5 medicines) (382, 95.5%) |
| 6 | 96 (24) |  |
| 7 | 35 (8.8) |  |
| 8 | 32 (8) |  |
| 9 | 3 (0.8) |  |
| 10 | 1 (0.2) | Severe polypharmacy ($\geq 10$ medications) (2, 0.5%) |
| 11 | 1 (0.2) |  |
| Total | 400 (100) |  |

**Table 4:** Total direct expenditure on medicines (USD) (Total expenditure by all 400 patients: USD 3409.54) (Mean± SD: 8.66±6.04) (Mean expenditure per medicine: 1.5)

| Expenditure (USD) | Frequency (%) |
| --- | --- |
| 1.01 - 11 | 314 (78.5) |
| 11.01 - 21 | 72 (18) |
| 21.01 - 31 | 11 (2.8) |
| 31.01 - 41 | 2 (0.5) |
| 61.01+ | 1 (0.3) |
| Total | 400 (100) |

The multinomial logistic regression analysis showed that the odd of a male suffering from moderate polypharmacy with respect to mild one was 1/2.33 = 0.43 time the odds for a female whereas it was 1/(9.82*10^32^) = 1.01*10^−33^ time in case of severe polypharmacy. There were significant impacts of all levels of education on both moderate and severe polypharmacy with reference to the mild one (except postgraduate for moderate only). Occupation-wise also, there were significant impacts of all occupations on both moderate and severe polypharmacy with reference to the mild one (except military, garment worker and retired worker for moderate only). Diagnosis-wise there were mixed types of significance for various disease conditions on both moderate and severe polypharmacy. Total cost of medications had no significant impact on both moderate and severe polypharmacy (p-values 0.443 and 0.393 respectively). (Table 5)

**Table 5:** Multinomial logistic regression of moderate and severe polypharmacy in reference to mild polypharmacy with various predictor variables (n = 400)

| Predictors | Moderate polypharmacy |  |  | Severe polypharmacy |  |  |
| --- | --- | --- | --- | --- | --- | --- |
|  | Beta (SE) | p-value | 95% CI for OR (lower & upper) | Beta (SE) | p-value | 95% CI for OR (lower & upper) |
| Intercept | 271.94 (1.33) | - | $1.27*10^{118}$<br>($9.25*10^{116}$ - $1.74*10^{119}$) | 8.31 (0.35) | | 4090.78 (2047.58- 8172.82) |
| Age | 0.007 (0.03) | 0.82 | 1.01 (0.94- 1.07) | -2.11 (25.9) | 0.934 | 0.12 ($1.08*10^{-23}$ - $1.34*10^{21}$) |
| Gender (Female) | 0.84 (1.88) | 0.654 | 2.33 (0.05- 9.45) | 75.96 (0.35) | <0.001 | $9.82*10^{32}$ ($4.91*10^{32}$ - $1.96*10^{33}$) |
| Education (Primary) | -37.06 ($1.2*10^{-9}$) | <0.001 | $7.96*10^{-17}$ ($7.96*10^{-17}$ - $7.96*10^{-17}$) | 57.25 ($1.2*10^{-9}$) | <0.001 | $7.36*10^{24}$ ($7.36*10^{24}$ - $7.36*10^{24}$) |
| Education (Secondary) | 15.83 ($6.14*10^{-9}$) | <0.001 | $7.55*10^6$ ($7.55*10^6$ - $7.55*10^6$) | 34.14 ($2.15*10^{-19}$) | <0.001 | $6.77*10^{14}$ ($6.77*10^{14}$ - $6.77*10^{14}$) |
| Education (Higher secondary) | -2.45 (1.05) | 0.02 | 0.08 (0.01- 0.68) | -106.81 (0.0003) | <0.001 | $4.07*10^{-47}$ ($4.06*10^{-47}$ - $4.07*10^{-47}$) |
| Education (Undergraduate) | -328.01 (0.61) | <0.001 | $3.5*10^{-143}$ ($1.05*10^{-143}$ - $1.16*10^{-142}$) | -169.27 ($4.05*10^{-24}$) | <0.001 | $3.05*10^{-74}$ ($3.05*10^{-74}$ - $3.05*10^{-74}$) |
| Education (Postgraduate) | -7.15 (NaN) | NaN | $7.79*10^{-4}$ (NaN - NaN) | 6.55 ($1.53*10^{-104}$) | <0.001 | 705.32 (705.32 – 705.32) |
| Occupation (Farmer) | -267.96 (0.92) | <0.001 | $4.22*10^{-117}$ ($6.9*10^{-118}$ - $2.58*10^{-116}$) | -168.15 ($8.47*10^{-16}$) | <0.001 | $9.31*10^{-74}$ ($9.31*10^{-74}$ - $9.31*10^{-74}$) |
| Occupation (Housewife) | -268.21 (1.46) | <0.001 | $3.26*10^{-117}$ ($1.85*10^{-118}$ - $5.75*10^{-116}$) | -116.55 (0.35) | <0.001 | $2.41*10^{-51}$ ($1.21*10^{-51}$ - $4.82*10^{-51}$) |
| Occupation (Student) | 57.53 (0.61) | <0.001 | $9.7*10^{24}$ ($2.92*10^{24}$ - $3.21*10^{25}$) | 37.36 ($4.05*10^{-24}$) | <0.001 | $1.68*10^{16}$ ($1.68*10^{16}$ - $1.68*10^{16}$) |
| Occupation (Businessman) | 238.69 ($8.12*10^{-16}$) | <0.001 | $4.62*10^{103}$ ($4.62*10^{103}$ - $4.62*10^{103}$) | -16.84 (0) | <0.001 | $4.82*10^{-8}$ ($4.82*10^{-8}$ - $4.82*10^{-8}$) |
| Occupation (Military) | 31.21 (NaN) | NaN | $3.59*10^{13}$ (NaN - NaN) | 7.12 (0) | <0.001 | 1248.2 (1248.2 – 1248.2) |
| Occupation (Garment worker) | 64.63 (NaN) | NaN | $1.18*10^{28}$ (NaN - NaN) | 13.92 (0) | <0.001 | $1.11*10^6$ ($1.11*10^6$ - $1.11*10^6$) |
| Occupation (Engineer) | 7.74 ($9*10^{-16}$) | <0.001 | 2318.63 (2318.63-2318.63) | 7.47 ($8.82*10^{-154}$) | <0.001 | 1767.23 (1767.23 – 1767.23) |
| Occupation (Shopkeeper) | 67.21 ($1.74*10^{-15}$) | <0.001 | $1.55*10^{29}$ ($1.55*10^{29}$ - $1.55*10^{29}$) | 13.24 (0) | <0.001 | $5.67*10^5$ ($5.67*10^5$ - $5.67*10^5$) |
| Occupation (Retired worker) | 369.72 (NaN) | NaN | $3.69*10^{160}$ (NaN – NaN) | -4.23 (0) | <0.001 | 0.01 (0.01 –0.01) |
| Dx (Common cold, cough, fever) | 145.7 ($1.54*10^{-36}$) | <0.001 | $1.9*10^{63}$ ($1.9*10^{63}$ - $1.9*10^{63}$) | 76.76 ($8.05*10^{-30}$) | <0.001 | $2.18*10^{33}$ ($2.18*10^{33}$ - $2.18*10^{33}$) |
| Dx (Constipation) | 96.18 ($6.75*10^{-18}$) | <0.001 | $5.93*10^{41}$ ($5.93*10^{41}$ - $5.93*10^{41}$) | 73.9($6.75*10^{-18}$) | <0.001 | $1.24*10^{32}$($1.24*10^{32}$ - $1.24*10^{32}$) |
| Dx (COPD) | -0.38 (1.24) | 0.759 | 0.68 (0.05- 7.79) | -45.59 ($2.15*10^{-19}$) | <0.001 | $1.57*10^{-20}$ ($1.57*10^{-20}$ - $1.57*10^{-20}$) |
| Dx (Cough, Tonsillitis, Headache) | 111.54 ($5.56*10^{-50}$) | <0.001 | $2.76*10^{48}$ ($2.76*10^{48}$ - $2.76*10^{48}$) | -0.56 ($1.62*10^{-117}$) | <0.001 | 0.57 (0.57 –0.57) |
| Dx (DM) | 43.85 ($2.05*10^{-14}$) | <0.001 | $1.11*10^{19}$ ($1.11*10^{19}$ - $1.11*10^{19}$) | 28.98 ($2.05*10^{-14}$) | <0.001 | $3.89*10^{12}$ ($3.89*10^{12}$ - $3.89*10^{12}$) |
| Dx (Ear problem) | 71.33 ($8.46*10^{-20}$) | <0.001 | $9.6*10^{30}$ ($9.6*10^{30}$ - $9.6*10^{30}$) | 47.14 ($8.46*10^{-20}$) | <0.001 | $2.97*10^{20}$ ($2.97*10^{20}$ - $2.97*10^{20}$) |
| Dx (Enteric fever) | -0.37 (1.61) | 0.814 | 0.68 (0.02- 1.62) | 69.17 (0.35) | <0.001 | $1.1*10^{30}$($5.53*10^{29}$ - $2.2*10^{30}$) |
| Dx (Epistaxis) | 37.42 ($1.42*10^{-24}$) | <0.001 | $1.78*10^{16}$ ($1.78*10^{16}$ - $1.78*10^{16}$) | 21.74 ($7.42*10^{-29}$) | <0.001 | $2.78*10^9$ ($2.78*10^9$ - $2.78*10^9$) |
| Dx (Fever, Cough) | -2.83 (1.6) | 0.077 | 0.05 (0.01 - 1.36) | -517.62 ($6.98*10^{-135}$) | <0.001 | $1.57*10^{-225}$ ($1.57*10^{-225}$ - $1.57*10^{-225}$) |
| Dx (Fever, Cough, Muscle pain) | -1.41 (1.62) | 0.382 | 0.24 (0.01- 5.82) | -88.58 ($9.74*10^{-64}$) | <0.001 | $3.38*10^{-39}$ ($3.38*10^{-39}$ - $3.38*10^{-39}$) |
| Dx (Fever, Headache, Joint pain) | 93.12 ($3.99*10^{-41}$) | <0.001 | $2.77*10^{40}$ ($2.77*10^{40}$ - $2.77*10^{40}$) | 17.88 ($2.33*10^{-45}$) | <0.001 | $5.86*10^7$ ($5.86*10^7$ - $5.86*10^7$) |
| Dx (Fracture, Pain, Weakness) | 57.79 ($2.31*10^{-26}$) | <0.001 | $1.25*10^{25}$ ($1.25*10^{25}$ - $1.25*10^{25}$) | 15.12 ($1.03*10^{-99}$) | <0.001 | $3.69*10^6$ ($3.69*10^6$ - $3.69*10^6$) |
| Dx (Fungal infection, Itching) | -0.1 (1.55) | 0.945 | 0.89 (0.04 – 19.1) | 86.87 (0.0003) | <0.001 | $5.33*10^{37}$ ($5.33*10^{37}$ - $5.34*10^{37}$) |
| Dx (Gastritis, N,V&D) | 134.34 ($8.85*10^{-59}$) | <0.001 | $2.22*10^{58}$ ($2.22*10^{58}$ - $2.22*10^{58}$) | 29.87 ($3.45*10^{-124}$) | <0.001 | $9.44*10^{12}$ ($9.44*10^{12}$ - $9.44*10^{12}$) |
| Dx (Backache, Bone pain) | -0.8 (1.64) | 0.625 | 0.44 (0.01 - 11.24) | -4.39 ($6.12*10^{-64}$) | <0.001 | 0.01 (0.01 –0.01) |
| Dx(Hyperlipidemia, HTN) | 38.66 (2.31*10<sup>-18</sup>) | <0.001 | 6.21*10<sup>16</sup> (6.21*10<sup>16</sup> - 6.21*10<sup>16</sup>) | 10.82 (1.84*10<sup>-121</sup>) | <0.001 | 50440.14 (50440.14 – 50440.14) |
| Dx (Hyperlipidemia, HTN, DM) | 133.57 (1.01*10<sup>-58</sup>) | <0.001 | 1.02*10<sup>58</sup> (1.02*10<sup>58</sup> - 1.02*10<sup>58</sup>) | 14.15 (1.52*10<sup>-138</sup>) | <0.001 | 1.4*10<sup>6</sup> (1.4*10<sup>6</sup> - 1.4*10<sup>6</sup>) |
| Dx (HTN, DM) | 33.06 (1.16*10<sup>-80</sup>) | <0.001 | 2.298*10<sup>14</sup> (2.02*10<sup>33</sup> - 2.02*10<sup>33</sup>) | 11.15 (1.16*10<sup>-80</sup>) | <0.001 | 70130.22 (70130.22 – 70130.22) |
| Dx (HTN, DM, Asthma) | 76.68 (1.2*10<sup>-9</sup>) | <0.001 | 2.02*10<sup>33</sup> (2.02*10<sup>33</sup> - 2.02*10<sup>33</sup>) | 87.32 (1.2*10<sup>-9</sup>) | <0.001 | 8.43*10<sup>37</sup> (8.43*10<sup>37</sup> - 8.43*10<sup>37</sup>) |
| Dx (HTN, Muscle pain) | 33.69 (1.42*10<sup>-159</sup>) | <0.001 | 4.27*10<sup>14</sup> (4.27*10<sup>14</sup> - 4.27*10<sup>14</sup>) | 6.71 (0) | <0.001 | 823.14 (823.14 – 823.14) |
| Dx (Hyperthyroidism) | -0.15 (1.63) | 0.923 | 0.85 (0.03 – 21.18) | 35.64 (6.16*10<sup>-42</sup>) | <0.001 | 3.01*10<sup>15</sup> (3.01*10<sup>15</sup> - 3.01*10<sup>15</sup>) |
| Dx (Hyperuricemia, Arthritis) | 48.05(6.08*10<sup>-21</sup>) | <0.001 | 7.37*10<sup>20</sup> (7.37*10<sup>20</sup> - 7.37*10<sup>20</sup>) | 83.71 (2.71*10<sup>-49</sup>) | <0.001 | 2.27*10<sup>36</sup> (2.27*10<sup>36</sup> - 2.27*10<sup>36</sup>) |
| Dx (Urolithiasis, Fever) | 122.82 (3.69*10<sup>-54</sup>) | <0.001 | 2.2*10<sup>53</sup> (2.2*10<sup>53</sup> - 2.2*10<sup>53</sup>) | 5.57 (1.75*10<sup>-118</sup>) | <0.001 | 262.9 (262.9 – 262.9) |
| Dx (Urolithiasis, Urine problem) | -0.37 (1.53) | 0.806 | 0.68 (0.03 – 13.91) | 35.02 (3.09*10<sup>-17</sup>) | <0.001 | 1.62*10<sup>15</sup> (1.62*10<sup>15</sup> - 1.62*10<sup>15</sup>) |
| Dx (Migraine headache) | 43.36 (4.24*10<sup>-20</sup>) | <0.001 | 6.82*10<sup>18</sup> (6.82*10<sup>18</sup> - 6.82*10<sup>18</sup>) | 24.56 (8.82*10<sup>-72</sup>) | <0.001 | 4.67*10<sup>10</sup> (4.67*10<sup>10</sup> - 4.67*10<sup>10</sup>) |
| Dx (Nerve pain) | 100.7 (9.44*10<sup>-45</sup>) | <0.001 | 5.46*10<sup>43</sup> (5.46*10<sup>43</sup> - 5.46*10<sup>43</sup>) | -9.09 (5.46*10<sup>-66</sup>) | <0.001 | 1.12*10<sup>-4</sup> (1.12*10<sup>-4</sup> - 1.12*10<sup>-4</sup>) |
| Dx (Parasitic infection) | -0.96 (1.32) | 0.465 | 0.38 (0.02 - 5.08) | -222.64 (2.07*10<sup>-84</sup>) | <0.001 | 2.01*10<sup>-97</sup> (2.01*10<sup>-97</sup> - 2.01*10<sup>-97</sup>) |
| Dx (Per vaginal bleeding, weakness) | -875.48 (0) | <0.001 | 0 | -307.85 (0) | <0.001 | 1.99*10<sup>-134</sup> (1.99*10<sup>-134</sup> - 1.99*10<sup>-134</sup>) |
| Dx (Piles, Constipation) | 189.52 (8.48*10<sup>-83</sup>) | <0.001 | 2.04*10<sup>82</sup> (2.04*10<sup>82</sup> - 2.04*10<sup>82</sup>) | 7.46 (0) | <0.001 | 1740.31 (1740.31 – 1740.31) |
| Dx (RTI) | 252.71 (8.47*10<sup>-16</sup>) | <0.001 | 5.67*10<sup>109</sup> (5.67*10<sup>109</sup> - 5.67*10<sup>109</sup>) | 421.47 (8.47*10<sup>-16</sup>) | <0.001 | 1.1*10<sup>183</sup> (1.1*10<sup>183</sup> - 1.107737*10<sup>183</sup>) |
| Dx (Scabies) | 18.22 (1.13*10<sup>-9</sup>) | <0.001 | 82449914.29 (8.24*10<sup>7</sup> - 8.24*10<sup>7</sup>) | 7.24 (9.3*10<sup>-94</sup>) | <0.001 | 1401.27 (1401.27 – 1401.27) |
| Dx (Sleeplessness, Anxiety) | 120.83 ($2.5 \times 10^{-53}$) | <0.001 | $2.99 \times 10^{52}$ ($2.99 \times 10^{52}$ - $2.99 \times 10^{52}$) | 3.5 ($4.05 \times 10^{-99}$) | <0.001 | 8.99 (8.99–8.99) |
| Dx (TB) | 188.33 ($3.13 \times 10^{-82}$) | <0.001 | $6.22 \times 10^{81}$ ($6.22 \times 10^{81}$ - $6.22 \times 10^{81}$) | 7.73 (0) | <0.001 | 2297.85 (2297.85 – 2297.85) |
| Total Cost of Medications (USD) | 0.03 (0.03) | 0.443 | 1.03 (0.95- 1.11) | 5.74 (6.72) | 0.393 | 312.57 ($5.87 \times 10^{-4}$ - $1.66 \times 10^8$) |
| Dx: Diagnosis; RTI: Respiratory tract infection; TB: tuberculosis; HTN: hypertension; DM: diabetes mellitus; mild polypharmacy: 1–4 medications; moderate polypharmacy: 5–9 medications; severe polypharmacy: $\geq 10$ medications<br>Model: $P(\text{Status of Polypharmacy}) = 1 / \{ 1 + e^{-(a + b1\text{Age} + b2\text{Gender} + b3\text{Education level} + b4\text{Occupation} + b5\text{Diagnosis} + b6\text{Total cost of medicines (USD)})} \}$ | | | | | | |

Pantoprazole was the most frequently prescribed medicine (177, 7.8%) and azithromycin (118, 5.2%) the commonest antibiotic prescribed. A combination of simeticone, aluminium hydroxide and sodium carboxymethyl cellulose was the commonest combination medicine (75, 3.3%) used. Total number of medicines consumed by all 400 patients was 2269, with mean±SD 5.67±1.08. (Annex 2)

## Discussions

Maximum patients (ie,382, 95.5%) experienced moderate polypharmacy (5-9 medications) in the present research. Both moderate and severe polypharmacy cases were non-significantly related with age, gender and total cost of medications. They had significant relationship in almost all levels of education and occupation and showed mixed type of significance and non-significance with the diagnosis of the respondents.A survey carried out in the USA showed the trend of polypharmacy among the patients aged 65 years as 42% (taking ≥5 medications), and 13% (taking ≥10 medications).^20^ Various researches also confirmed that the prevalence of polypharmacy increases with advancing age.^3,19,21^ Maanen et al. reported the majority of polypharmacy cases among patients aged > 70 years.^11^ However, Jansen and Brouwersreported the reduced incidence of polypharmacy among the frail community-dwellers aged > 85 years due to the underuse of medications for chronic diseases.^6^ But Hajjar et al. found that polypharmacy was in the increasing trend in the elderly, especially among aged > 85 years.^1^ Rozenfeld et al. reported that 37.2% of women and 25.8% of men used ≥5 medicines in Brazil.^12^

The studies conducted in different countries such as Saudi Arabia^22^,Oman^23^, Brazil^24^, and England^25^ also showed that gender had no significant impact on polypharmacy. However, research conducted in Kuwait showed significant relation of gender with polypharmacy.^26^ Such discrepancy in the output with the demographic characteristics might be one of the beauties of epidemiologic research with a diverse population. The study also reported that education beyond primary education significantly decreased polypharmacy as the case in the present study i.e, the lower the education, the higher was the incidence of polypharmacy.^22,26-28^ Salih et al. reported significant relationship of eight different diagnoses such as dyslipidemia, hypertension, diabetes mellitus, osteoarthritis, bronchial asthma, osteoporosis, heart failure and coronary artery disease with the polypharmacy.^22^ Similarly, the study by Al-Hashar et al. also reported that polypharmacy was significantly correlated with the number of comorbidities as well as with the cardiovascular disease as the admitting diagnosis.^23^ Mukete and Ferdinandfound that the hypertensive adults were more likely to be the victims of polypharmacy, and the associated complications namely increased risk of adverse events (fall injury, hyperkalemia and hypokalemia, heart failure, and blood pressure exacerbation), drug-drug interaction, and augmented costs. They reported that the use of non-steroidal anti-inflammatory drugs (NSAIDs) by older people might increase the risk of antihypertensive medication initiation by 1.5 to 1.8 times.The estimated direct and indirect costs of treatment of hypertension might rise to USD274 billion by 2030 from USD46.4 billion in 2010. Control in polypharmacy could avoid USD900 million to USD 1.7 billion unwanted expenses in medicines.^29^ Hajjar et al. reported that patients taking > 2 psychotropic agents might be in 2.4 to 4.5-fold increased risk of falling.^1^

Rozenfeld et al. analyzed both the quantitative and qualitative implications of polypharmacy and found that polypharmacy cannot be totally avoided in every instancebecause of the advancing diseases such as hypertension and diabetes mellitus (highly prevalent among the elderly), requiring necessity for polypharmacy.^12^ Rational prescribing and dispensing is the essence of cost-effective health care asithelps reduce health care burden and future potential complications. The prescribing indicators developed by the WHO in collaboration with the International Network for Rational Use of Drugs (INRUD) can be used to detect and solve prescribing problems, including polypharmacy.^30^ Both prescription and non-prescription medications (and even those brought by the patients during all health care visits for review) should be considered during detection of polypharmacy cases.^1,12^ The Beers Criteria was developed to explore a list of potentially harmful medications in the community-dwelling elderly.^31^ Unnecessary use of medicines can be reduced by adhering to the guidelines; avoiding prescribing to relieve the common self-limiting ailments; regular medication reviews; prescription audits; and with the computerized alert systems.^12^

The present research explored that pantoprazole was the commonest medicine (177, 7.8%), and azithromycin (118, 5.2%) the commonest antibiotic prescribed. A combination of simeticone, aluminum hydroxide and sodium carboxymethyl cellulose was the commonest combination medicine (75, 3.3%) used. Hajjar et al. reported that the most frequently prescribed medications were estrogen products, levothyroxine, hydrochlorothiazide, atorvastatin, and lisinopril.Most widely used non-prescription medications were pain medications (acetaminophen, ibuprofen, acetylsalicylic acid), cold and cough medications (eg, pseudoephedrine, diphenhydramine), and vitamin or nutrient products (eg, multivitamins, vitamins E and C, ginseng, Ginkgo biloba extract).^1^ The data collected by National Health and Nutrition Examination Surveys (NHANES) from 1988-1994, 1999-2004 and 2005-2010 confirmed that an increased number of antihypertensives were used by older Americans. They explored the reasons and found that advancing age was leading to a decline in hypertension control.^32^ Guaraldo et al. stated that the practical alterations in all pharmacokinetic processes(absorption, first-pass metabolism,bioavailability, distribution, protein building, renal andhepatic clearance) in the elderly patients are the commonly experienced phenomena in their therapy.^33^

Mismanagement of polypharmacy in community and hospital pharmacies may contribute to ADR development and drug-drug interaction in patients of all age groups, especially children and the elderly. Pharmacists’ interventions on irrational and avoidable polypharmacy minimization can improve patients’ economic, clinical and humanistic outcomes (ECHO) in health care settings.^34,35^ Responsible use of medicines can eliminate at least USD213 billion by addressing six key areas namely non-adherence, delayed evidence-based therapy, antibiotic misuse, medication errors, suboptimal generic use, and mismanaged polypharmacy.^20^

## Strengths and limitations of this study

- The present study was one of the largest community-based researches in Nepal to explore polypharmacy cases and costs associated with polypharmacy.
- Still, thefew communities might not represent the whole country and the whole population.
- Generalizability (external validity) on polypharmcy issues may be somehow limited in the wider arena.
- Nevertheless, it might serve as a beacon to explore the polypharmacy cases in the similar settings. Prospective cohort studies could be conducted later based on the findings of the present study.

## Conclusions

Polypharmacy has two domains: appropriate and avoidable; the former can be continued as such as the latter is to be prevented as far as possible. Both moderate and severe polypharmacy cases were non-significantly related with age, gender and total cost of medications. They had significant relationship in almost all levels of education and occupation and showed mixed type of significance and non-significance with the diagnosis of the respondents. Polypharmacy cases can be minimized, taking into consideration the ADRs and drug interactions. Further studies are warranted in medication utilization and avoidable polypharmacy along with the detailed pharmacoeconomic evaluation (i.e., cost-effectiveness, cost-utility analysis) of the same.

## Data Availability

All data supporting the findings of this study are contained within the manuscript. Any additional information regarding the study including the questionnaires would be shared by the corresponding author upon request.

## Acknowledgements

The authors are grateful to Pokhara University Research Center (PURC) for providing the research grant (PURC 1/2073/74) to carry out the present research. They are also very much grateful to all the respondents for their valuable time and active cooperation throughout the research. The authors would like to acknowledge Dr. Sunil Shrestha, PharmD from Nepal Health Research and Innovation Foundation (NHRIF) for his thorough English language editing of the manuscript.

## Declarations

### Ethics approval and consent to participate

The study was ethically approved by the Nobel College Institutional Review Committee, Sinamangal, Kathmandu (NIRC 0103/2017).

## Consent for publication

Not applicable

## Authors’ Contributions

All authors made substantial contributions to conception and design, acquisition of data, or analysis and interpretation of data; took part in drafting the article or revising it critically for important intellectual content; gave final approval of the version to be published; and agree to be accountable for all aspects of the work.

